# Vascular smooth muscle calcium sparks and sarcoplasmic reticulum calcium load are reduced in women with preeclampsia

**DOI:** 10.1101/2025.11.04.25339536

**Authors:** Luisa C Parnell, Anna L Tierney, Elizabeth C Cottrell, Karolina Krakowiak, Richard D Unwin, Harry AT Pritchard, Alison M Gurney, Jenny E Myers, Adam Greenstein, Stephanie A Worton

## Abstract

**Background:** Maternal microvascular dysfunction is a hallmark of the hypertensive pregnancy disorder preeclampsia, but the mechanism(s) have not been defined. Historically, attention has focused on the vascular endothelium, but microvascular function is largely determined by the downstream effector, vascular smooth muscle cells (VSMCs). Within VSMCs, the principal pressure-induced vasodilatory pathway depends upon stimulation of large-conductance Ca^2+^-activated potassium channels (BK_Ca_) by localised Ca^2+^-release events from sarcoplasmic reticulum (SR) ryanodine receptors (‘Ca^2+^ sparks’). Here, Ca^2+^ sparks are assessed in preeclampsia for the first time.

**Methods:** Pressurised omental resistance arteries from normotensive pregnant women and women with preeclampsia were imaged by high-speed spinning-disk laser confocal microscopy to assess Ca^2+^ sparks (20-120 mmHg) and caffeine-evoked Ca^2+^ release (10 mmol/L; 80 mmHg). A liquid chromatography–selected reaction monitoring–mass spectrometry (LC-SRM-MS) assay was developed to measure SR Ca^2+^ pump Sarcoplasmic/Endoplasmic Reticulum Ca^2+^ ATPase 2 (SERCA2) and unphosphorylated/phosphorylated proteoforms of regulatory phospholamban. Vasoactive effects of the BK_Ca_–dependent vasorelaxant Human β-Defensin-2 (HBD2; 0.01-10 nmol/L) were assessed by wire myography.

**Results:** Ca^2+^ spark frequency was decreased at all intraluminal pressures in women with preeclampsia compared to normotensive pregnancy (p=1.1×10^-10^). In preeclampsia there was reduced SR Ca^2+^ released in response to caffeine (p=5.1×10^-4^) and the ratio of SERCA2 to inhibitory unphosphorylated phospholamban was decreased (p=0.030), indicating increased SERCA2 inhibition. In arteries from women with preeclampsia, HBD2 (0.01-10 nmol/L) caused BK_Ca_-dependent relaxation (p=4.6×10^-10^) without affecting Ca^2+^ spark frequency.

**Conclusions:** In microvascular arteries from women with preeclampsia, there is a striking reduction in Ca^2+^ spark frequency, likely secondary to increased inhibition of SERCA2 by unphosphorylated phospholamban and reduced SR Ca^2+^ load. These defects may contribute to vascular dysfunction in preeclampsia and present novel therapeutic targets to restore physiological control of the vasculature. Meanwhile, BK_Ca_-mediated relaxation by HBD2 independent of Ca^2+^ sparks supports vasodilatory strategies bypassing disrupted microvascular mechanisms in preeclampsia.

## Introduction

Preeclampsia, affecting 2-8% of pregnancies, remains a leading cause of global maternal mortality and morbidity^1^ and contributes to 10-25% of global stillbirths^2^. Whilst the origins of preeclampsia lie in the stressed placenta, the maternal syndrome is characterised by widespread vascular dysfunction which manifests as increased systemic vascular resistance, hypertension, reduced perfusion, and subsequent end-organ dysfunction^3^. Women with preeclampsia have consistently demonstrated endothelial dysfunction^4^, which despite being poorly defined is widely accepted to be a hallmark of the disease. Although isolated arteries from multiple vascular beds demonstrate increased vasoconstriction and reduced endothelium-dependent relaxation, reports of the endothelial pathways affected are inconsistent and specific mechanisms of microvascular dysfunction have not been defined^5–11^. Whilst clinical management of preeclampsia aims to ameliorate complications of the disorder, no treatments target the underlying pathophysiology, and delivery of the fetus remains the only cure. Poorly-controlled maternal hypertension is the leading indication for preterm delivery in preeclampsia^12^, and preeclampsia contributes to approximately 13.5% of births before 34 weeks in the UK^13^. A new therapeutic approach predicated on identifying and targeting the specific cellular mechanisms of microvascular dysfunction is urgently needed.

In microvascular arteries, vascular smooth muscle cells (VSMCs) are the downstream effectors controlling arterial lumen diameter and are therefore the key determinants of flow. In response to intraluminal pressure, global increases in VSMC Ca^2+^ cause vasocontraction, but there is simultaneous activation of vasorelaxatory pathways, the balance of which determines myogenic tone^14,15^. The principal vasodilatory response to intraluminal pressure is the release of spatiotemporally regulated Ca^2+^ events, termed Ca^2+^ sparks, from ryanodine receptors (RyRs) on the sarcoplasmic reticulum (SR) membrane^15–17^. Ca^2+^ sparks are large-amplitude, highly-localised increases in cytosolic [Ca^2+^], which activate ∼30 nearby large-conductance potassium-activated Ca^2+^ channels (BK_Ca_ channels) on the VSMC membrane, resulting in spontaneous transient outward currents (STOCs), membrane hyperpolarisation and vasorelaxation^15^. Pressure-induced activation of Ca^2+^ sparks is mediated via the production of oxidants which activate Protein Kinase G1α (PKG1α) through a mechanism distinct from activation by cGMP^18^. Whilst it has not been delineated in vascular smooth muscle, in cardiac myocytes oxidant–activated PKG1α regulates Ca^2+^ sparks through either a direct interaction with the RyR or indirectly via Ser16 phosphorylation of phospholamban (PLN) which relieves inhibition of the Sarcoplasmic / Endoplasmic Reticulum Ca^2+^ ATPase 2 (SERCA2) pump^18–22^. SERCA2 activity determines the SR [Ca^2+^], which directly affects the open probability of RyRs^23^.

Identifying new therapeutics that can cause vasorelaxation through BK_Ca_ activation is expected to be advantageous for the treatment of hypertension. Human β-defensin-2 (HBD2), a naturally-occurring defensin peptide of the innate immune system, causes vasorelaxation and reduces blood pressure (BP) in normotensive rats and monkeys, reportedly via direct BK_Ca_ activation^24^.

We previously determined that the vasorelaxant effects of the tryptophan metabolite kynurenine are mediated via the Ca^2+^ spark–BK_Ca_ vasoregulatory axis^25^. In arteries from women with preeclampsia, kynurenine remains a potent vasorelaxant, but its effects are reduced compared with arteries from normotensive women, which may indicate an underlying perturbation in this pathway^25^. This hypothesis is supported by observations in the preeclampsia-like model of hypoxic pregnant sheep, in which hypoxia abrogates the normal pregnancy-induced increase in Ca^2+^ spark and STOC frequency observed in normoxic pregnant sheep^26,27^. However, Ca^2+^ sparks were studied in a denuded, unpressurised, *en face* arterial preparation^26,27^, which does not account for the pressure sensitivity of this vasodilatory pathway.

Here, we present the first investigation of Ca^2+^ sparks in human preeclampsia, and their regulation by SR Ca^2+^ load and PLN phosphorylation. We identify a substantial reduction in Ca^2+^ spark frequency in microvascular arteries from women with preeclampsia, which is secondary to a decreased ratio of SERCA2 to unphosphorylated PLN, causing increased SERCA2 inhibition and reduction of total SR Ca^2+^ load. We demonstrate that activation of BK_Ca_ channels by HBD2, independent of Ca^2+^ sparks, causes relaxation of arteries from women with preeclampsia.

## Methods

Detailed methods can be found in the Data Supplement. Please see the Major Resources Table in the Supplemental Materials.

In brief, omental biopsies were obtained with informed consent at the time of caesarean section from normotensive women with uncomplicated pregnancies and women with a clinical diagnosis of preeclampsia. In pressurised omental resistance arteries, Ca^2+^ sparks were imaged using high-speed spinning-disk confocal microscopy. Ca^2+^ release events were measured at pressures of 20, 60, 80 and 110 mmHg, or in response to sequential caffeine boluses (10 mmol/L), or following 30 minutes incubation with HBD2 (10 nmol/L).

Snap-frozen omental arteries were lysed and trypsin digested. LC-SRM-MS analysis was performed using an Agilent 6495 triple quadrupole mass spectrometer to monitor PLN and SERCA2 peptides, including phosphorylated PLN isoforms. Relative ratio of SERCA2 to both phosphorylated and unphosphorylated phospholamban isoforms was determined using heavy isotope-labelled internal standard peptides.

Relaxation of preconstricted omental arteries (U46619 EC_80_) mounted on the wire myograph was assessed in response to vehicle or HBD2 (0.001-10 nmol/L) in the presence or absence of BK_Ca_ inhibitors (paxilline 1.5 µmol/L or iberiotoxin 100 nmol/L). Constriction in response to U46619 (10^-10^–10^-5.7^ mol/L) was assessed before and after 1 hour incubation with HBD2 (10 nmol/L) or vehicle.

## Results

### Participants

The characteristics of 86 normotensive pregnant women and 27 women with preeclampsia included in this study are shown in Table 1. As expected, all women with preeclampsia had hypertension requiring antenatal antihypertensive treatment, they were more likely to be nulliparous and the gestational age at delivery and offspring birthweight centile were significantly reduced. Of note, 20/27 women delivered before 34 weeks. Forty-four percent of women with preeclampsia received intravenous magnesium sulphate immediately prior to delivery, indicated for severe maternal disease, fetal neuroprotection or both. Representative subgroups of participants contributed to each experiment, and within each subgroup there remained no significant difference in maternal characteristics.

**Table 1:**
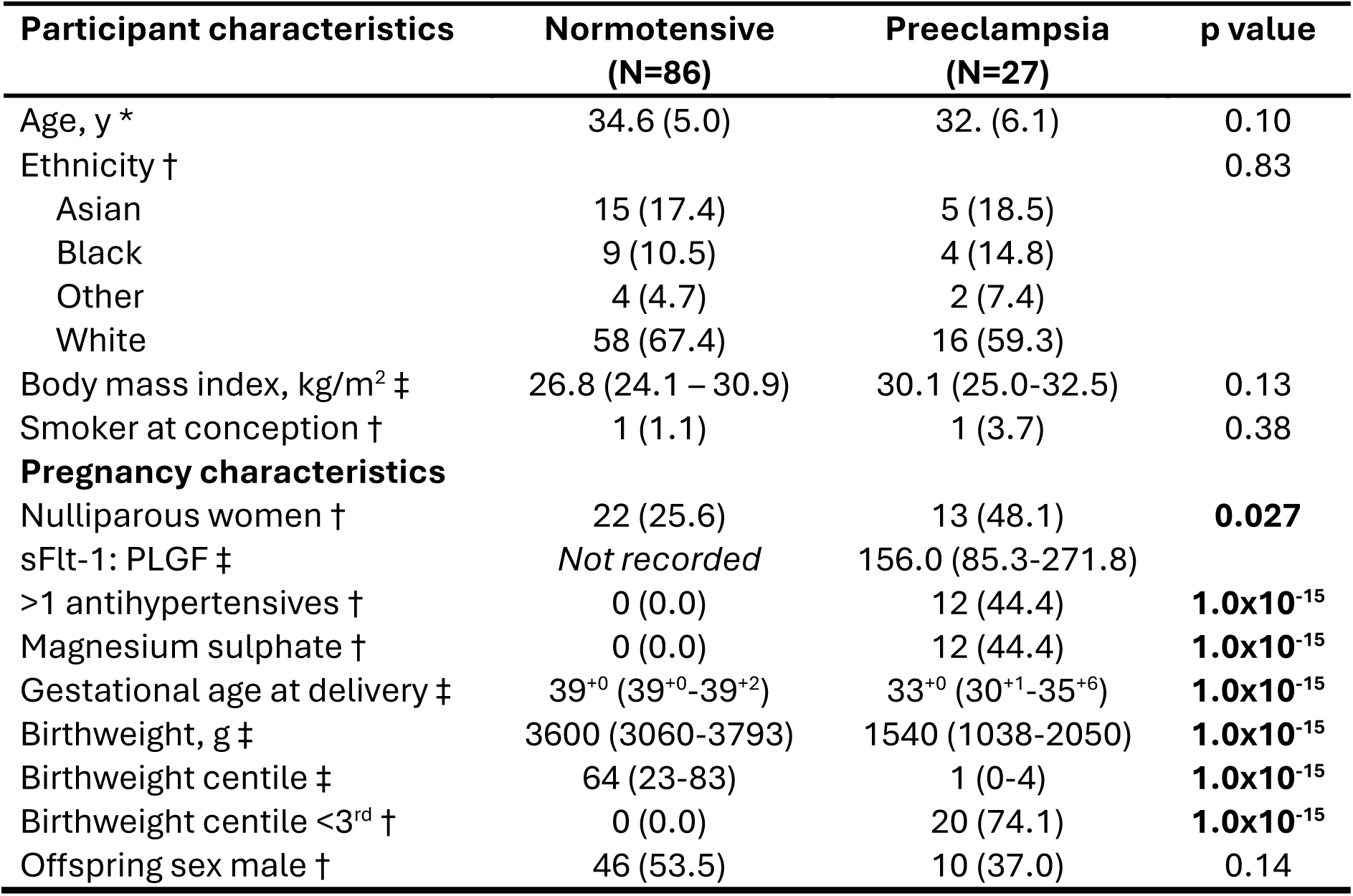
Characteristics of participants. * Data shown as mean (SD); compared with unpaired t-test. † Data shown as n (%); compared with chi-squared test. ‡ Data shown as median (IQR); compared with Mann-Whitney test.

| Participant characteristics | Normotensive<br>(N=86) | Preeclampsia<br>(N=27) | p value |
| --- | --- | --- | --- |
| Age, y * | 34.6 (5.0) | 32. (6.1) | 0.10 |
| Ethnicity † |  |  | 0.83 |
| Asian | 15 (17.4) | 5 (18.5) |  |
| Black | 9 (10.5) | 4 (14.8) |  |
| Other | 4 (4.7) | 2 (7.4) |  |
| White | 58 (67.4) | 16 (59.3) |  |
| Body mass index, kg/m<sup>2</sup> ‡ | 26.8 (24.1 – 30.9) | 30.1 (25.0-32.5) | 0.13 |
| Smoker at conception † | 1 (1.1) | 1 (3.7) | 0.38 |
| **Pregnancy characteristics** |  |  |  |
| Nulliparous women † | 22 (25.6) | 13 (48.1) | **0.027** |
| sFlt-1: PLGF ‡ | *Not recorded* | 156.0 (85.3-271.8) |  |
| >1 antihypertensives † | 0 (0.0) | 12 (44.4) | **1.0x10<sup>-15</sup>** |
| Magnesium sulphate † | 0 (0.0) | 12 (44.4) | **1.0x10<sup>-15</sup>** |
| Gestational age at delivery ‡ | 39<sup>+0</sup> (39<sup>+0</sup>-39<sup>+2</sup>) | 33<sup>+0</sup> (30<sup>+1</sup>-35<sup>+6</sup>) | **1.0x10<sup>-15</sup>** |
| Birthweight, g ‡ | 3600 (3060-3793) | 1540 (1038-2050) | **1.0x10<sup>-15</sup>** |
| Birthweight centile ‡ | 64 (23-83) | 1 (0-4) | **1.0x10<sup>-15</sup>** |
| Birthweight centile <3<sup>rd</sup> † | 0 (0.0) | 20 (74.1) | **1.0x10<sup>-15</sup>** |
| Offspring sex male † | 46 (53.5) | 10 (37.0) | 0.14 |

### Ca^2+^ sparks are reduced in omental arteries from women with preeclampsia

In pressurised omental arteries from normotensive pregnant women, Ca^2+^ spark frequency was low at low intraluminal pressures, and as expected, increased with rising intraluminal pressure to peak at physiological pressure of 80 mmHg (Figure 1A+B). Ca^2+^ spark frequency was significantly reduced in arteries from women with preeclampsia compared to normotensive women across the full range of pressures tested (preeclampsia effect p=1.1×10^-10^; Figure 1A+B). Ca^2+^ spark amplitude, duration, rise time and decay time did not differ between pregnancy groups (Figure 1C-F).

**Figure 1:**
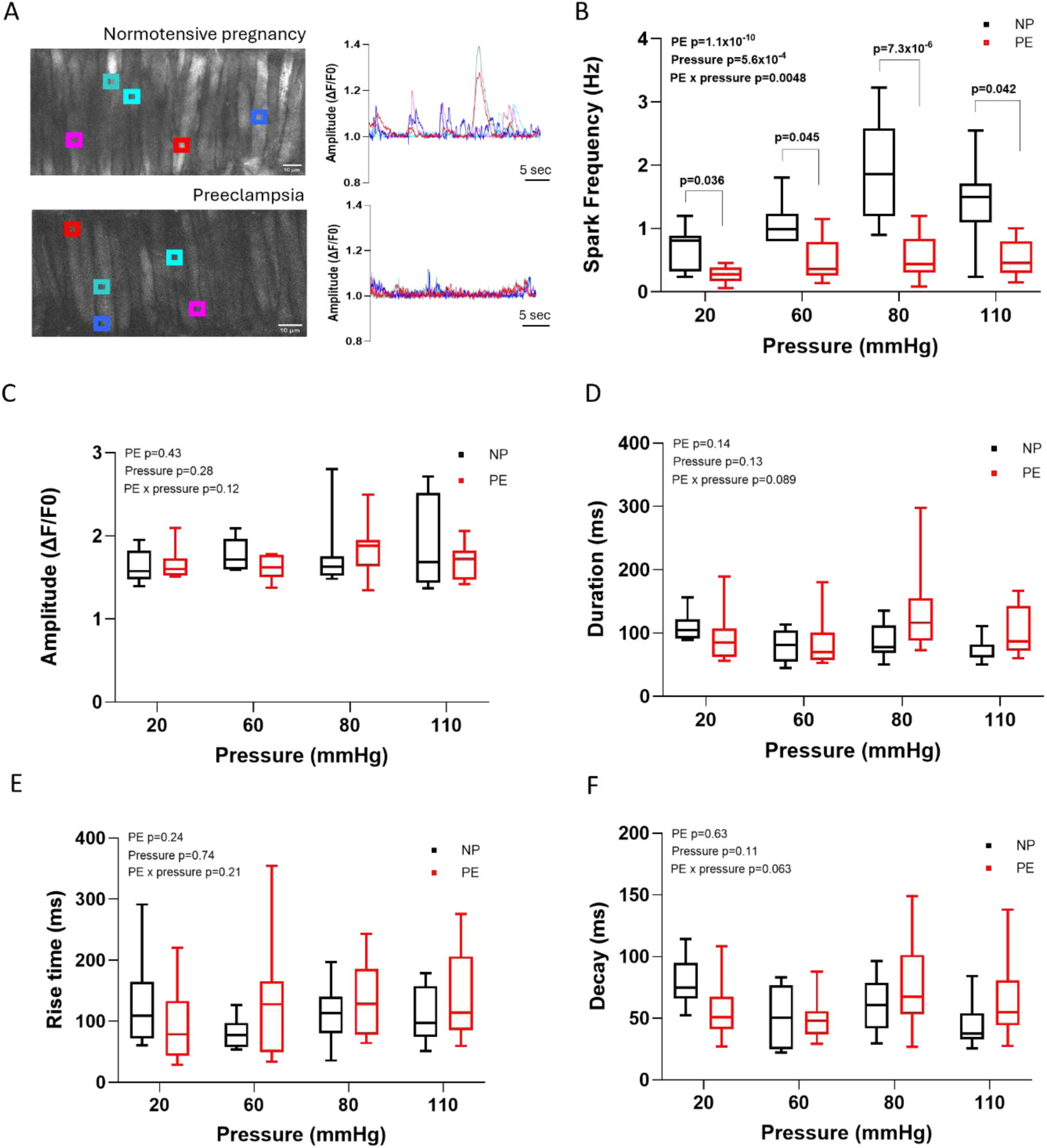
Ca^2+^ spark frequency is reduced across a range of pressures in omental resistance arteries from women with preeclampsia. Arteries incubated with Ca^2+^ fluorescent dye Cal-520-AM and imaged using high-speed spinning-disk laser confocal microscopy at increasing intraluminal pressures (20, 60, 80 and 110 mmHg). **A**: Representative images (left) of arteries from women with normotensive pregnancy (NP) or preeclampsia (PE) with regions of interest (coloured boxes) identified by SparkAN with corresponding coloured lines (right) showing change in fractional fluorescence (F/F0). **B-F:** Ca^2+^ spark frequency (**B**), amplitude (**C**), duration (**D**), rise time (**E**) and decay time (**F**) were compared between women with normotensive pregnancy (NP, black; N=7-15) and preeclampsia (PE, red; N=8-13). Data expressed as median with range. NP and PE groups compared using mixed-effects analysis with Sidak’s multiple comparison tests performed at each pressure. All p values indicated on graphs.

To determine any effect of maternal medications, Ca^2+^ spark frequency at 80 mmHg was analysed according to the antihypertensive regime (no antihypertensive, labetalol only, nifedipine only or amlodipine), number of antihypertensive agents, or administration of magnesium sulphate; there were no differences in Ca^2+^ spark frequency related to maternal medications (Data supplement Figure S-V A-C). Linear regression was used to study the association between Ca^2+^ spark frequency and maternal characteristics that could potentially affect Ca^2+^ spark frequency (maternal age^28^ and maternal BMI^14^) or surrogate markers of disease severity (gestational age at delivery, birthweight z score, or maternal serum biomarkers; Data supplement Table S-II). After adjusting for preeclampsia diagnosis, there was no association between Ca^2+^ spark frequency and maternal characteristics (age or BMI) or pregnancy outcomes (gestation at delivery or birthweight z score). Maternal serum biomarkers (soluble FMS-like tyrosine kinase (sFlt-1), placental growth factor (PlGF), or sFlt-1:PlGF)) with adjustment for the test-to-delivery interval, were not associated with Ca^2+^ spark frequency amongst women with preeclampsia.

### Caffeine-evoked Ca^2+^ transients are reduced in omental arteries from women with preeclampsia

The open probability of RyRs is determined by the SR [Ca2+]^23^. Therefore, having identified reduced frequency of Ca^2+^ spark release in preeclampsia, we proceeded to assess the total Ca^2+^ store within the SR by the application of 10 mmol/L caffeine which sensitises RyRs to Ca^2+^ causing release of Ca^2+^ stores^20^ (Figure 2A+B). For caffeine-evoked Ca^2+^ release, area under curve (AUC) and peak amplitude were significantly reduced in arteries from women with preeclampsia compared to normotensive pregnancy (Figure 2B-D). Duration and rise were unchanged between groups. In arteries from women with preeclampsia, there was a slower decay in normalised Ca^2+^ transient curves.

**Figure 2:**
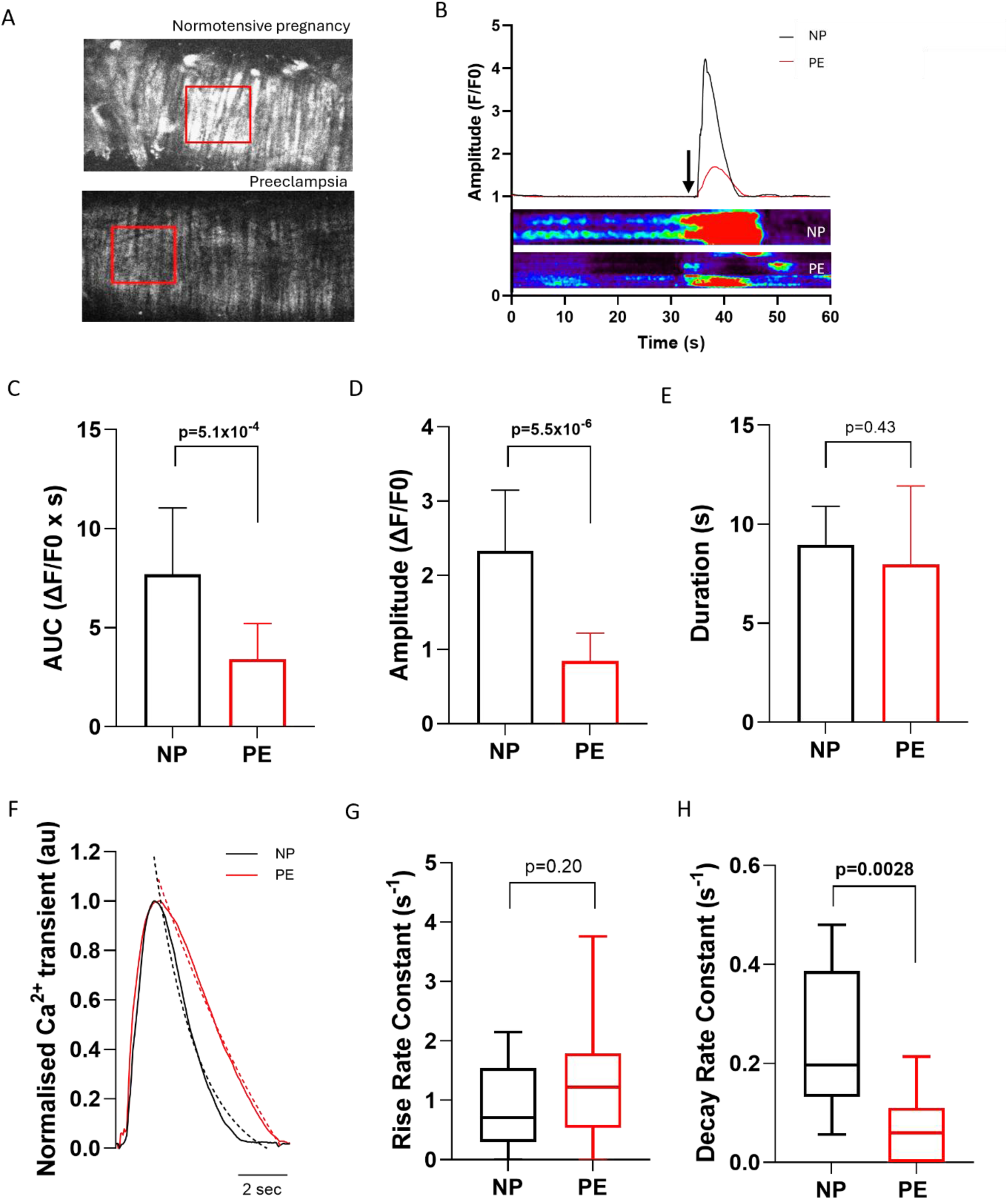
Sarcoplasmic reticulum Ca^2+^ stores are reduced in omental resistance arteries from women with preeclampsia. Omental arteries incubated with Ca^2+^ fluorescent dye Cal-520-AM were pressurised to 80 mmHg and imaged using high-speed spinning-disk confocal microscopy. Caffeine (10 mmol/L) was applied to arteries, and transient Ca^2+^ release measured using SparkAN software. **A**: Representative images of a region of interest (ROI) placed over a section of an artery. **B**: Representative trace and visualisation of the change in fluorescence compared to baseline fluorescence (F/F0) in arteries from women with normotensive pregnancy and preeclampsia. Arrow indicates addition of caffeine bolus. **C-H**: Kinetics of transient Ca^2+^ release following addition of a caffeine bolus. Area under curve (AUC; **C**), amplitude (**D**), and duration (**E**) in arteries from women with normotensive pregnancy (NP, N=12) and women with preeclampsia (PE, N=13). (**F-H**): Caffeine-evoked Ca^2+^ release peaks were normalised (peak maximum=1; representative peaks shown in **F**), and rate constant compared between women with normotensive pregnancy (N=11-12) and women with preeclampsia (N=10-12) for rise (**G**) and decay (**H**), with **F** illustrating a representative normalised curve of caffeine-evoked Ca^2+^ release for each group. Data presented as mean ±SD and compared with unpaired t-tests (**C-E**) or presented as median with range and compared by Mann-Whitney tests (**G+H**). Au refers to arbitrary units (**F**). All p values indicated on graphs.

Ca^2+^ ‘reuptake’ into the SR was assessed in a subset of arteries by application of a second caffeine bolus 2 minutes after the first. AUC, peak amplitude and decay remained reduced in the preeclampsia group following the second caffeine exposure. Duration and rise constant remained unchanged between groups (Data supplement Figure S-VI A-E).

There were no differences in caffeine-evoked Ca^2+^ transient AUC related to maternal antihypertensive regime, number of antihypertensive agents, or administration of magnesium sulphate (Data supplement Figure S-V D-F). After adjusting for preeclampsia diagnosis, there was no association between caffeine-evoked Ca^2+^ transient AUC and maternal BMI, gestation at delivery or birthweight z score (Data supplement Table S-III). Amongst women with preeclampsia, maternal serum biomarkers, adjusted for the test-to-delivery interval, were not significantly associated with caffeine-evoked transient AUC (Data supplement Table S-III). There was a trend towards a reduction in AUC with increasing maternal age (β=-0.017, 95% CI [-0.038, 0.0028]; p=0.088; Data supplement Table-III + Figure S-VIIA) and decreasing Log(PlGF) (β=0.74, 95% CI [-0.043, 1.52]; p=0.061; Data supplement Table S-III and Figure S-VII-B), although the strength of these associations is uncertain.

### SERCA2:PLN is decreased in omental arteries from women with preeclampsia

It was hypothesised that reduced caffeine-evoked Ca^2+^ release in women with preeclampsia could be caused by reduced SR Ca^2+^ uptake by the SERCA2 pump due to inhibition by unphosphorylated PLN. Abundance of SERCA2 and phospholamban proteoforms were assessed by reference to heavy-labelled standard peptides in lysates from microvascular arteries using LC-SRM-MS. In women with preeclampsia, there was a significant decrease in the ratio of the SERCA2 to inhibitory unphosphorylated PLN (Figure 3), indicating greater inhibition of the SERCA2 pump in preeclampsia. Our custom proteomic assay differentiated between unphosphorylated PLN and its differentially phosphorylated proteoforms (Ser16, Thr17, Ser16/Thr17; Data supplement Figure S-I-III), but the abundance of phosphorylated PLN proteoforms in lysates of unpressurised arteries was expectedly low and below the limits of detection of the instrument (360 pmol/L of lysate).

**Figure 3:**
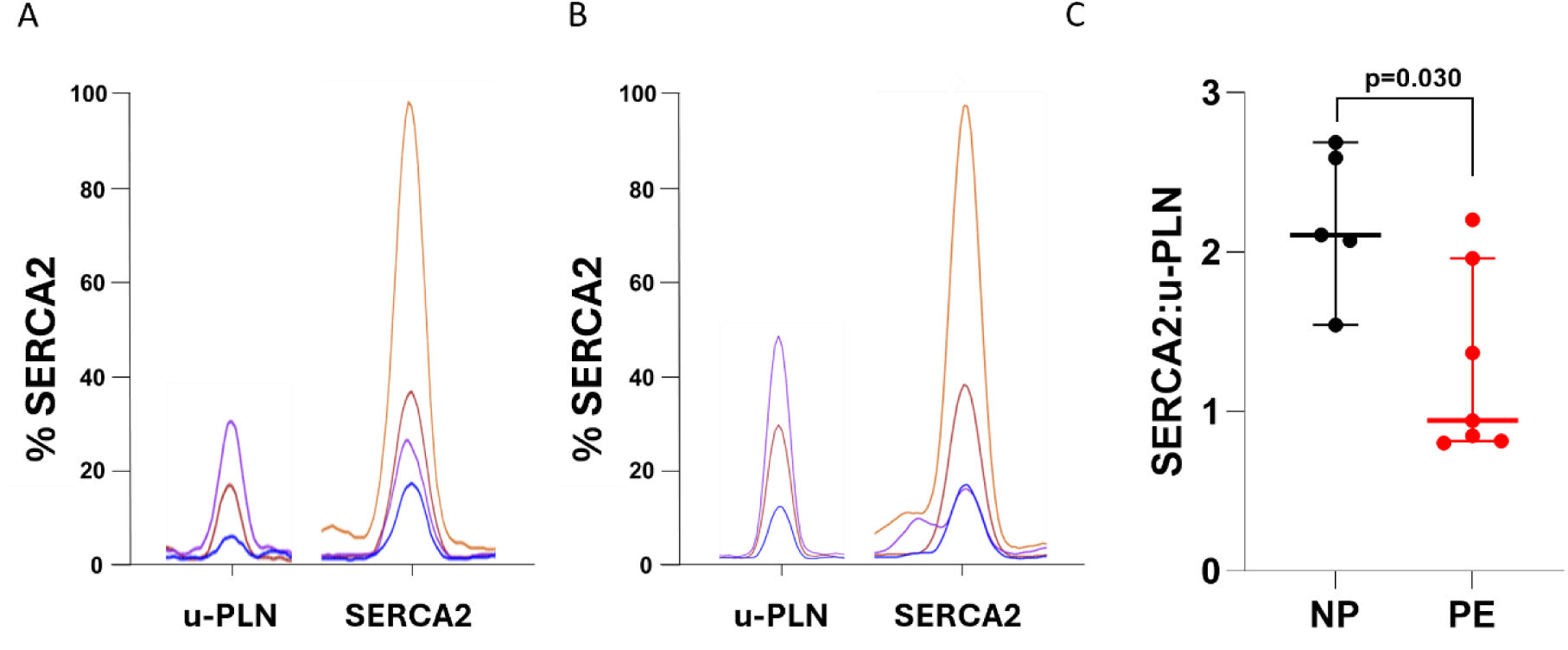
SERCA2:unphosphorylated PLN ratio is decreased in omental resistance arteries from women with preeclampsia. LC-SRM-MS assessment of unphosphorylated PLN (u-PLN) and SERCA2. **A**+**B**: Representative extracted ion chromatograms of u-PLN-specific and SERCA2-specific peptides in arteries from women with normotensive pregnancy (**A**) or preeclampsia (**B**). **C**: Ratio of SERCA2:u-PLN in arteries from women with normotensive pregnancy or preeclampsia. Due to small sample sizes, individual data points are shown with median indicated; N=5-7. Ratios compared between groups using Mann-Whitney test, p value indicated on graph.

### HBD2 relaxes sub-maximally contracted omental arteries from women with normotensive pregnancy and preeclampsia, independent of Ca^2+^ sparks

Having established a defect in the Ca^2+^ spark pathway in preeclampsia, we proceeded to explore the therapeutic potential of bypassing Ca^2+^sparks to activate BK_Ca_ channels directly in women with preeclampsia, using the putative antihypertensive HBD2. In omental arteries mounted on the wire myograph (normotensive mean diameter 323.6±58.5 μm, preeclampsia mean diameter 328.8±48.2 μm), endothelium-dependent relaxation of U46619-constricted arteries to bradykinin (10^-5^ mol/L) was reduced in arteries from women with preeclampsia compared to those from normotensive pregnant women (preeclampsia median relaxation 26.4 % [IQR 8.8 - 60.1] vs. normotensive 85.6 % [57.3 - 92.6]; p=1.0×10^-15^). In arteries sub-maximally contracted (EC_80_) with U46619, HBD2 (0.01-10 nmol/L) induced significant vasorelaxation compared to vehicle control in arteries from normotensive pregnant women (Figure 4A+B) and women with preeclampsia (Figure 4C). Direct comparison of concentration-relaxation curves for HBD2 demonstrated no difference between normotensive women and women with preeclampsia (RM 2-way ANOVA, preeclampsia effect p=0.24; graph not shown).

**Figure 4:**
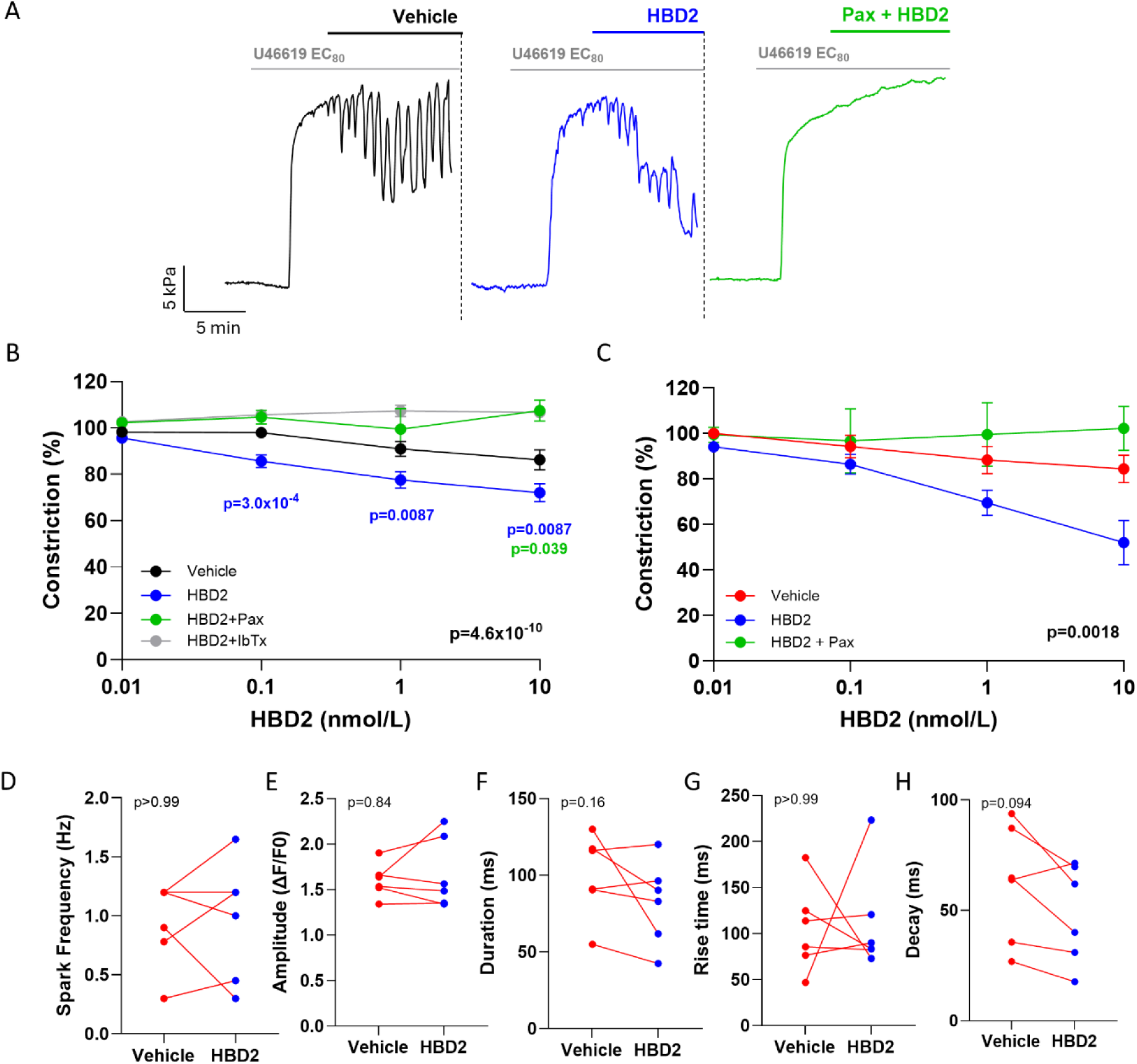
HBD2 causes BK_Ca_-mediated vasorelaxation of pre-constricted omental resistance arteries. **A**: Representative wire myography traces of omental arteries from a normotensive pregnant woman submaximally pre-constricted with U46619 (EC_80_) and treated with HBD2 (0.01-10 nmol/L; blue) or vehicle (physiological saline solution [PSS]; black) in the presence or absence of paxilline (Pax, 1.5 µmol/L; orange). **B**: Concentration-relaxation curves for pre-constricted omental arteries from normotensive pregnancy treated with vehicle (PSS; black), HBD2 only (0.01-10 nmol/L; blue), HBD2 + paxilline (1.5 µmol/L; green) or HBD2 + iberiotoxin (IbTx, 100 nmol/L; grey); N=7-30, n=14-44. **C**: Concentration-relaxation curves for pre-constricted omental arteries from women with preeclampsia treated with vehicle (PSS; red), HBD2 only (0.01-10 nmol/L; blue) or HBD2 + paxilline (1.5 µmol/L; green); N=8, n=14-20. Data expressed as mean±SEM; compared using RM mixed effects analysis (**B**) or 2-way ANOVA (**C**) with Sidak’s multiple comparison tests to vehicle-treated arteries. **D-H:** Ca^2+^ spark kinetics in pressurised (80 mmHg) omental arteries from women with preeclampsia (N=6, n=12) following treatment with HBD2 (10 nmol/L; blue) or vehicle control (PSS; red); Ca^2+^ spark frequency (**D**), amplitude (**E**), duration (**F**), rise time (**G**) and decay time (**H**). Individual data points shown, compared by Wilcoxon matched-pairs signed-rank test. Individual p values indicated on graphs.

HBD2 was confirmed to act via a BK_Ca_-dependent mechanism in a subset of experiments by inhibition of HBD2-induced relaxation with BK_Ca_ channel inhibitors paxilline (1.5 μmol/L) or iberiotoxin (100 nmol/L; NP group only; Figures 4B+C). It was noted that BK_Ca_ inhibitors also abolished the characteristic oscillations of sub-maximally constricted omental arteries as previously described^25^. There were no other underlying differences in arterial diameter, baseline or post-treatment constriction to 120 mmol/L KPSS or maximal constriction to U46619 (10^-5.7^ mol/L) either between arteries allocated to each treatment group or between arteries from normotensive women compared with arteries from women with preeclampsia (data not shown).

To determine if HBD2-induced relaxation was mediated by an effect on Ca^2+^ sparks, Ca^2+^ sparks were assessed in pressurised omental arteries from women with preeclampsia treated with HBD2 (10 nmol/L, 30 minutes). HBD2 did not increase Ca^2+^ spark frequency, nor were there statistically significant differences in any other Ca^2+^ spark kinetics compared to untreated arteries (Figure 4D-H).

### HBD2 reduces U46619-induced constriction in omental arteries from women with normotensive pregnancy and women with preeclampsia

To further assess vasoactive effects of HBD2, U46619-induced constriction (10^-10^-10^-5.7^ mol/L) was measured before and after 1-hour incubation with HBD2 (10 nmol/L) or vehicle control (Figure 5A). In omental arteries from normotensive pregnant women and women with preeclampsia, HBD2 significantly attenuated constriction in response to U46619 (Figure 5B+C). This effect of HBD2 on U46619-induced vasoconstriction did not persist following washout of HBD2 or vehicle (Figure 5D), and HBD2 treatment had no effect on subsequent maximal constriction to 120 mmol KPSS. There were no underlying differences in diameter, pre-treatment constriction to 120 mmol KPSS or pretreatment constriction to U46619 either between arteries allocated to each treatment group or between arteries from normotensive pregnancy or preeclampsia (data not shown).

**Figure 5:**
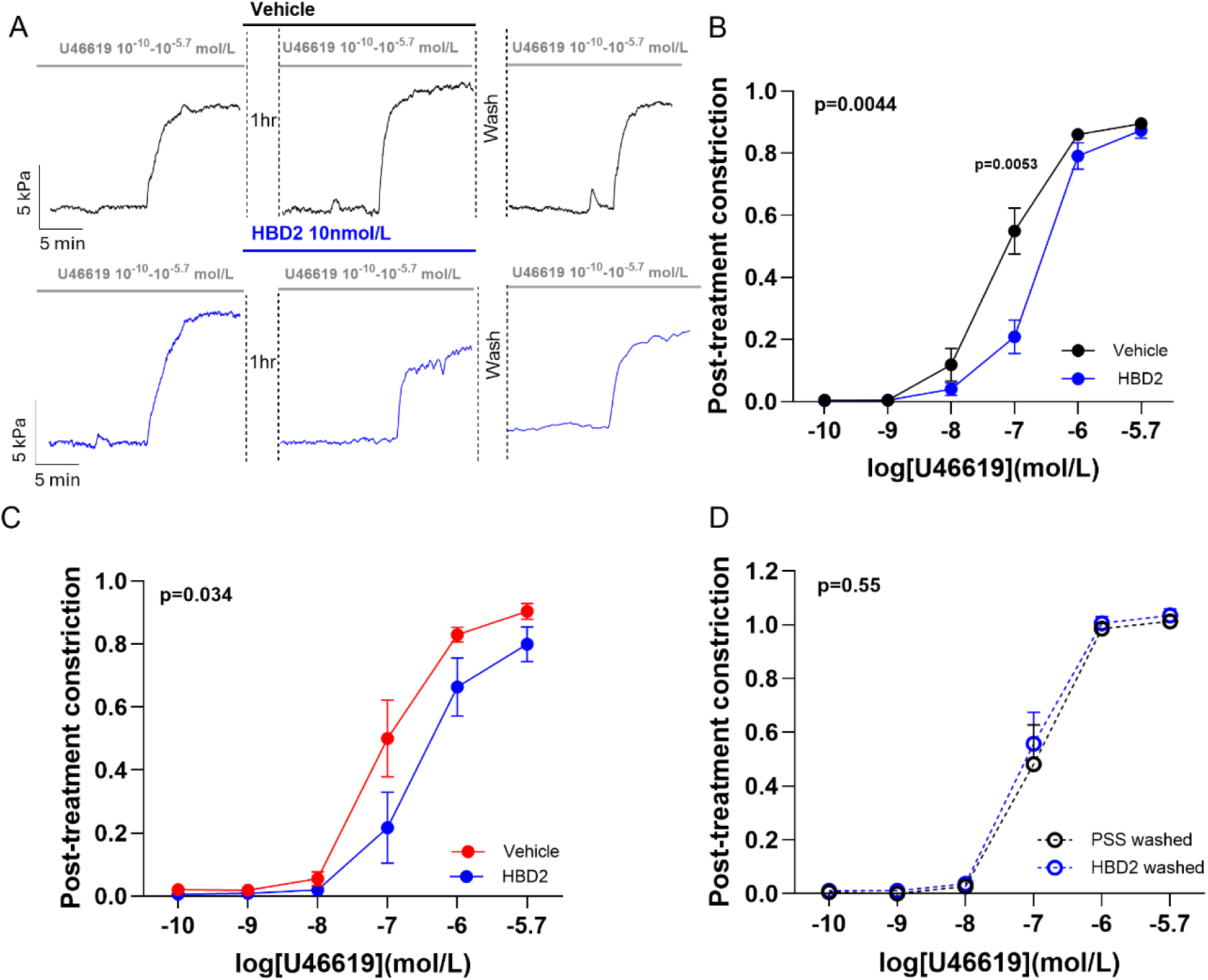
HBD2 treatment reduces vasoconstriction in omental resistance arteries from normotensive women and women with preeclampsia. **A:** Representative wire myography traces of omental arteries from normotensive pregnant women constricted with U46619 (10^-10^-10^-5.7^mol/L) before and after 1 hour incubation with HBD2 (10 nmol/L; blue) or vehicle (physiological saline solution [PSS]; black), and following washout of HBD2 or vehicle. **B:** Post-treatment constriction expressed relative to maximal pre-treatment constriction to U46619 (10^-5.7^ mol/L) following 1 hour incubation with HBD2 (10 nmol/L; blue) or vehicle (PSS; black) in arteries from normotensive pregnant women. N=15. **C:** Post-treatment constriction expressed relative to maximal pre-treatment constriction to U46619 (10^-5.7^ mol/L) following 1 hour incubation with HBD2 (10 nmol/L; blue) or vehicle (PSS; red) in arteries from women with preeclampsia. N=8. **D:** Post-washout constriction expressed relative to maximal pre-treatment constriction to U46619 (10^-5.7^ mol/L) following washout of HBD2 (10 nmol/L; blue, dashed line) or vehicle (PSS; black, dashed line) in arteries from normotensive pregnant women. N=9. Data expressed as mean±SEM and compared using RM ANOVA with Sidak’s multiple comparison tests. All overall p values shown on graphs.

## Discussion

For the first time, data presented here demonstrate differences in SR Ca^2+^ handling in the VSMCs of intact microvascular arteries from a well-characterised cohort of women with preeclampsia. There are three key findings from this study. Firstly, the frequency of pressure-induced Ca^2+^ sparks released from the SR is markedly reduced in preeclampsia. Secondly, we have shown that the total SR Ca^2+^ store is reduced in preeclampsia and that there is a decrease in amounts of SERCA2 relative to its inhibitor, unphosphorylated PLN. Finally, we have shown that the purported novel antihypertensive HBD2 can cause vasorelaxation and reduce vasoconstriction in human microvasculature arteries, an effect which is preserved in the hypertensive pathology preeclampsia.

The fine-tuning of microvascular tone in response to changes in intraluminal pressure by Ca^2+^ sparks is one of the key determinants of vascular tone and has been extensively established^15,16,18,21^. Here, the relationship between intraluminal pressure and Ca^2+^ sparks is confirmed for the first time in omental arteries from normotensive pregnant women. The increase in Ca^2+^ spark frequency in response to pressure was similar to that described in mouse mesenteric arteries^16^; albeit that in normotensive women, maximal spark activation occurred at 80 mmHg with a small decline at the highest pressure tested (110 mmHg), compared with a plateau of maximal Ca^2+^ sparks ≥50 mmHg in mice^16^. Interpretation of findings is limited by the paucity of existing data in human arteries and the traditional neglect of studying arteries from females and specifically during pregnancy.

In women with preeclampsia, Ca^2+^ spark frequency was globally reduced across all pressures, but the difference between groups was greatest at physiological intraluminal pressure (80 mmHg), where there was a 75% reduction. Whilst we have not assessed the downstream effects of this reduction in Ca^2+^ sparks, reduced Ca^2+^ spark frequency or any other mechanism interrupting the spatiotemporal coupling of sparks to BK_Ca_ channels is universally associated with a reduction in STOCs and increased arterial constriction^14,15,29,30^, which would contribute to the vascular dysfunction of preeclampsia. In animal studies, an isolated reduction in VSMC Ca^2+^ spark frequency without changes in amplitude or Ca^2+^ spark kinetics has been reported in models of pre-eclampsia, obesity, Alzheimer’s disease, and subarachnoid haemorrhage^14,27,30,31^. In the only one of these studies where Ca^2+^ sparks were assessed at multiple pressures^14^, obese and lean mice had comparable Ca^2+^ sparks at 20 mmHg, whilst in preeclampsia we have observed a reduction at 20 mmHg and a failure to increase with changes in pressure. In the hypoxic sheep model of preeclampsia, which exhibits hypertension and increased uterine vascular resistance, Ca^2+^ spark frequency in uterine arteries was measured in unpressurised, *en face* arteries, which makes comparison with our findings difficult^27^. In this animal model, Ca^2+^ spark frequency increased in normoxic animals in pregnancy^26^, whilst in hypoxic sheep, Ca^2+^ sparks failed to increase in pregnancy and remained similar to non-pregnant animals^27^. Reference to a non-pregnant female comparator group, which was not possible in our study, would provide useful context to our findings to indicate whether differences in Ca^2+^ spark frequency at 20 mmHg might be contributed to by a failed adaptation to pregnancy in women with preeclampsia.

SR Ca^2+^ load, which is a key physiological regulator of Ca^2+^ spark frequency via a direct effect on the open state probability of RyRs^23^, is shown here to be substantially reduced in preeclampsia. Within VSMCs, SR Ca^2+^ stores are principally determined by the active uptake of cytosolic Ca^2+^ by SERCA2 and its release via RyR and inositol 1,4,5-triphosphate receptors. In preeclampsia, reduced caffeine-evoked Ca^2+^ transient AUC and slower reuptake of Ca^2+^ into the SR implicate SERCA2 in the mechanism of reduced Ca^2+^ sparks^32^. In our experiments, repeated caffeine boluses assessed 2 minutes apart yielded comparable results, indicating that SERCA2 is consistently reloading the SR with a reduced SR [Ca^2+^] in preeclampsia.

SERCA2 activity, which controls the SR Ca^2+^ load and thereby Ca^2+^ spark frequency^32,33^, is inhibited by PLN. Phosphorylation of PLN by PKG1α relieves this inhibition of SERCA2 activity^22,34^. To test the hypothesis that decreased SR Ca^2+^ load in arteries from women with preeclampsia could be due to inhibition of SERCA2 by unphosphorylated PLN, we developed a bespoke LC-SRM-MS assay to allow quantitation of SERCA2 and PLN proteoforms within microvascular arteries with comparable diameter to those used for live-cell Ca^2+^ imaging experiments. This approach confers significant advantages over traditional approaches such as Western blotting, due to the small tissue volumes required to assess multiple proteins and the ability to definitively identify proteins, distinguish between phosphoforms, and assign estimated concentrations by comparison to heavy-labelled standards. By developing the assay to reliably distinguish between differentially phosphorylated forms of PLN, we can state with certainty that the PLN reported is the unphosphorylated form. In VSMCs, PLN expression is much lower than cardiac myocytes^34^ and the phosphorylated proportion (Ser16, Thr17 or Ser16/Thr17 phosphorylated) in unpressurised arteries expectedly fell below the limit of detection of this method. In arteries from women with preeclampsia, the ratio of SERCA2 to inhibitory unphosphorylated PLN is decreased, indicating greater inhibition of SERCA2 which likely contributes to the reduction in SR Ca^2+^ load. This reduction in SERCA2:PLN ratio could follow from perturbations in thus far unexplored earlier steps of this pathway where pressure induces oxidant generation and activation of PKG1α; this is discussed further below in relation to potential therapeutic interventions. Whilst our key findings of reduced caffeine-evoked Ca^2+^ transients and decreased SERCA2:PLN ratio create a compelling story to explain the mechanisms of reduced Ca^2+^ spark frequency in preeclampsia, we acknowledge that this does not exclude a contribution by other mechanisms that can affect Ca^2+^ spark frequency such as: altered RyR expression, clustering or sensitivity^32,33,35^; Ca^2+^ spark initiation by lysosomal TRPML1^36^; or Ca^2+^ spark boosting by extracellular Ca^2+37^.

Findings presented here indicate substantial alterations in SR Ca^2+^ handling in the VSMCs of arteries from women with preeclampsia, which challenges the accepted understanding of vascular dysfunction in preeclampsia as being principally driven by endothelial cell dysfunction. The role of the vascular smooth muscle (VSM) in preeclampsia was under consideration over 50 years ago when the enhanced vasopressor response to angiotensin II was reported to precede the onset of clinical disease^38^. Subsequently, in isolated arteries, altered VSM morphology (increased media-to-lumen ratio)^39^ and increased Ca^2+^ sensitivity were reported^40,41^.

Our evidence of an impaired VSM vasodilatory mechanism in preeclampsia represents a significant advance in our understanding of vascular dysfunction in this disease. It will take substantial experimental effort to determine to what extent the defects observed in VSMC Ca^2+^ handling are (i) a downstream effect of endothelial dysfunction, (ii) parallel but unrelated to other mechanistic perturbations in the endothelium, or (iii) the predominant functional vascular defect. Certainly, there is extensive evidence of endothelial activation, altered endothelial function and impaired endothelium-dependent vasorelaxation in small arteries from women with pre-eclampsia^6,7,42^. However, it is worth noting that whilst impairments in endothelium-mediated relaxation are reported in arteries from multiple vascular beds in preeclampsia^5,6,9^, conflicting findings are reported between different groups, vascular beds and techniques^8,10,11^. In obesity-related hypertension, another condition traditionally ascribed to endothelial dysfunction, detailed interrogation of vasorelaxatory pathways determined that differences in Ca^2+^ spark activation rather than endothelial mechanisms were responsible for vascular dysfunction^14^.

The use of arteries from a single vascular bed is an important consideration of this study. Our interpretation of the results assumes that observations of altered Ca^2+^ handling in preeclampsia are not limited to the omentum. Vascular dysfunction in preeclampsia is systemic with all maternal organs affected, and as it is accepted to be secondary to a factor in the maternal circulation it must be supposed that all vascular beds are exposed to the same causative factors. Nevertheless, potential local vascular bed variations in the pathways studied makes systemic effects of vascular defects and the effect of therapeutics on these pathways harder to predict.

The primary aim of studying vascular dysfunction in preeclampsia is to identify new targets for treatments that can reduce the risk of severe hypertension and safely allow gestation to be extended. This work identifies two strategies for future investigation of new therapeutics in preeclampsia which could be applied singly or together; (1) to bypass Ca^2+^ sparks with agents such as HBD2, and (2) to repair disruptions in Ca^2+^ sparks. HBD2, postulated as a novel antihypertensive agent^24^, causes BK_Ca_-dependent increases in mesenteric flow in mice and reduced BP in Sprague-Dawley rats and rhesus monkeys^24^. Here, we assessed the vasoactive properties of HBD2 in human arteries for the first time, confirming that HBD2 reduces U46619-mediated constriction and induces relaxation in human arteries from normotensive women and those with preeclampsia. It is usually a limitation to assess the Ca^2+^ spark-BK_Ca_ pathway in the unpressurised wire myograph system because the absence of physiological pressure may cause experiments to overestimate the pharmacological effects of BK_Ca_ activation. However, we have demonstrated that pressure-induced activation of Ca^2+^ sparks is absent in arteries from women with preeclampsia, therefore vasorelaxatory effects seen on the wire myograph are anticipted to persist in a physiological system. The effects of HBD2 on maternal arteries are independent of Ca^2+^ sparks but BK_Ca_-dependent, which is consistent with HBD2s reported mechanism in HEK293 cells to increase BK_Ca_ single-channel currents and STOC frequency^24^.

The relaxatory effects of HBD2 support the potential of bypassing dysfunctional vascular pathways to cause BK_Ca_-mediated relaxation of microvascular arteries in preeclampsia, as previously demonstrated with kynurenine and NS11021^25^. If the vasorelaxant effects of HBD2 are extrapolated to other vascular beds, our findings suggest HBD2 may have potential for systemic effects on maternal BP in women with preeclampsia. Translating these findings towards the clinical application of HBD2 as an antihypertensive in preeclampsia would require evidence of systemic BP effects in pregnant whole-animal models and extensive safety data from pregnant animal models and human models of placental function. Although no such safety data currently exists, HBD2’s role as a naturally occurring defensin peptide that is heavily expressed in the reproductive tract^43^ and excreted in high levels in human breast milk^44^ are initially encouraging for a favourable safety profile.

It is anticipated that further defining underlying mechanisms of vascular dysfunction in preeclampsia will facilitate the development of treatments targeted to repair these mechanisms and restore physiological control of the vasculature. In preeclampsia, failure to increase Ca^2+^ sparks in response to increased intraluminal pressure could indicate an upstream failure of the mechanism by which VSMCs detect or respond to pressure. Under physiological conditions, pressure in VSMCs or stretch in myocytes cause the release of oxidants that provide a mechanism for fine-tuning the smooth muscle response to physical stimuli^18,22^. The pattern of reduced Ca^2+^ spark frequency, reduced SR Ca^2+^ load and increased inhibition of SERCA2 by unphosphorylated PLN observed in preeclampsia is replicated in cardiac myocytes from the PKG[C42S]KI knock-in transgenic mouse, which is resistant to oxidant C42 dimerization of PKG1α^22^. In this model, reduced VSMC Ca^2+^ sparks are associated with decreased BK_Ca_ channel activation, vasoconstriction and hypertension^18^; the pregnancy phenotype of this model has not yet been reported. Studying this upstream pathway in preeclampsia may allow us to further pinpoint the molecular cause of vascular dysfunction in preeclampsia. A defect at this stage of the pathway would be particularly encouraging for future intervention because the synthetic compound G1 targets this mechanism specifically by inducing oxidation of PKG1α at its C42 residue, causing a reduction in BP in hypertensive mice^45^.

The mechanism by which preeclampsia causes a defect in pressure-induced Ca^2+^ sparks has not been addressed by this work. We are confident that reduced Ca^2+^ sparks are not a global response to altered haemodynamics of hypertension, as evidenced by preservation of VSMC Ca^2+^ sparks in arteries from hypertensive BPH/2 mice and spontaneously hypertensive rats^29,46^. Preeclampsia is accepted to be dependent upon the presence of a placenta which releases circulating factors that induce the maternal phenotype^3,47^. Our working hypothesis is that a placental-derived circulating factor(s) interacts with the microvasculature to cause changes in the pathway regulating pressure-activated Ca^2+^ sparks. In preeclampsia, there is commonly rapid resolution of the hypertensive phenotype in the weeks following delivery, suggesting at least partial resolution of the vascular defect after pregnancy. However, the later return of hypertension and persistence of long-term cardiovascular risk amongst women after preeclampsia may indicate a persistent defect or vulnerability in this mechanism. Of note, perturbations in the Ca^2+^ spark – BK_Ca_ regulatory axis have been identified in the cerebral arteries of mouse models of vascular dementia and Alzheimer’s^29,30^, therefore a persistent defect or susceptibility within this pathway could explain the increased risk of vascular dementia amongst women after preeclampsia^48^.

The unavoidable increases in primiparity, prematurity, fetal growth restriction (FGR), and antihypertensive treatment in women with preeclampsia reflects the expected characteristics of this group. Clinical guidelines advocate delivery of women with preeclampsia after 37^+0^ weeks of pregnancy or earlier if there are features of maternal or fetal compromise^49^. In our cohort 20/27 women delivered before 34 weeks indicating a relatively severe pre-eclampsia phenotype, which represents those women and babies with the most to gain from advances in therapeutics. A pathology-free comparator group requiring preterm delivery by caesarean section does not exist for the comparative study of Ca^2+^ sparks at earlier gestations. However, observational studies suggest that vascular resistance plateaus in the third trimester^50^, and it is unlikely that further gestational changes in myogenic tone would contribute substantially to the results presented. Indeed, in linear regression models, gestational age did not have any independent effect on Ca^2+^ sparks or caffeine-induced Ca^2+^ transients. We considered that Ca^2+^ channel blockers such as nifedipine or amlodipine may reduce global intracellular Ca^2+^ and therefore contribute to reduced SR Ca^2+^ stores. However, we found no indication that different groups of antihypertensives, nor treatment with magnesium sulphate prior to birth, influenced Ca^2+^ measurements. We explored the relationship between our measurements of Ca^2+^ handling and those maternal characteristics that may be anticipated to affect the Ca^2+^ spark regulatory axis (age and BMI) and surrogate markers of disease severity, with no significant findings. There was a trend towards a 2% reduction in the size of the caffeine-evoked Ca^2+^ transient for each year of increased maternal age. Whilst this trend is in keeping with other studies demonstrating a decline in SERCA2 function with age^28^, this did not reach statistical significance. Maternal circulating levels of sFlt-1 and PlGF represent pathological and physiological factors released from the placenta respectively, and their ratio was used in this cohort of women with preeclampsia to aid diagnosis^49^. After adjusting for the test-to-delivery interval, there was a trend towards an association between caffeine-evoked Ca^2+^ transients and maternal serum PlGF, although not sFlt-1:PlGF ratio. Whilst it is biologically plausible that reduced PlGF could cause downstream changes in Ca^2+^ handling via a thus far unrecognised effect, low PlGF is frequently present in FGR without vascular dysfunction^51^, making it more likely that any possible association between PlGF and caffeine-induced Ca^2+^ transients reflects a shared dependence on placental failure rather than a direct causal relationship.

In conclusion, our data provide novel evidence of a defect in vasorelaxatory pathways in the VSM in preeclampsia, which builds upon widely-accepted evidence of endothelial dysfunction. In microvascular arteries from women with preeclampsia there are VSMC defects in SR Ca^2+^ handling and a reduction in Ca^2+^ spark frequency. Bypassing Ca^2+^ sparks to cause vasorelaxation, or therapeutically targeting the dysfunctional Ca^2+^ spark pathway, both offer strategies to treat vascular dysfunction in preeclampsia.

## Data Availability

All data is available upon reasonable request to the corresponding author

## Non-Standard Abbreviations and Acronyms

AU: Arbitrary Unit
AUC: Area Under Curve
BK_Ca_ Channel: Large-conductance Ca^2+^-activated Potassium Channel
BP: Blood Pressure
FGR: Fetal Growth Restriction
HBD2: Human β-Defensin-2
LC-SRM-MS: Liquid Chromatography–Selected Reaction Monitoring–Tandem Mass Spectrometry
NP: Normotensive Pregnancy
PKG1α: Protein Kinase G1α
PlGF: Placental Growth Factor
PLN: Phospholamban
RyR: Ryanodine Receptor
SERCA2: Sarcoplasmic/Endoplasmic Reticulum Ca^2+^ ATPase 2
sFlt-1: Soluble FMS-like Tyrosine Kinase
SR: Sarcoplasmic Reticulum
STOC: Spontaneous Transient Outward Current
VSM: Vascular Smooth Muscle
VSMC: Vascular Smooth Muscle Cell

## Acknowledgments

None.

## Supplemental Material

Supplemental Methods

Tables SI – SIII

Figure SI – SVII

Graphical Abstract

Supplemental References 52-57

## Novelty and Significance

### What is known?

- Vascular dysfunction is a key feature of the pregnancy-specific hypertensive disorder preeclampsia
- Activation of large-conductance Ca^2+^-activated potassium channels (BK_Ca_ channels) by ryanodine receptor-mediated release of Ca^2+^ sparks from the sarcoplasmic reticulum (SR) is a key determinant of myogenic tone

### What new information does this article contribute?

- In this first assessment of Ca^2+^ sparks in preeclampsia, we demonstrate that Ca^2+^ spark frequency is reduced in omental arteries from women with preeclampsia
- Ca^2+^ spark frequency is reduced due to reduced SR Ca^2+^ load, which is likely secondary to increased inhibition of SERCA2 by unphosphorylated phospholamban
- The BK_Ca_ channel activator Human β-Defensin 2 causes vasorelaxation of omental resistance arteries from normotensive pregnant women and women with preeclampsia

Preeclampsia is considered a disorder of endothelial dysfunction, but the mechanism(s) of vascular dysfunction have not been defined. In vascular smooth muscle, activation of BK_Ca_ channels by Ca^2+^ sparks released from the SR is the key mechanism regulating arterial relaxation in response to increases in pressure. Here, we study Ca^2+^ sparks in preeclampsia for the first time and show substantially decreased Ca^2+^ spark frequency in omental arteries from women with preeclampsia compared to normotensive pregnant women at a range of pressures. Reduced Ca^2+^ spark frequency is explained by reduced SR Ca^2+^ load in preeclampsia. Reuptake of Ca^2+^ into the SR is slower in preeclampsia, which may be explained by the decreased ratio of the SR Ca^2+^ pump, SERCA2, to its inhibitor unphosphorylated phospholamban. We provide the first direct evidence of a defect in smooth muscle vasorelaxatory pathways in preeclampsia. Thus, we identify potential new mechanisms for treating preeclampsia, either through strategies to restore Ca^2+^ spark frequency or to bypass Ca^2+^ sparks to activate BK_Ca_ channels directly. In support of this, we demonstrate that the BK_Ca_ activator Human β-Defensin 2, a purported antihypertensive, causes vasorelaxation in human arteries for the first time and this effect is preserved in preeclampsia.

